# The Full Database of Countries with Potential COVID-19 Data Misreport based on Benford’s Law

**DOI:** 10.1101/2020.12.04.20243832

**Authors:** Ahmad Kilani, Georgios P. Georgiou

## Abstract

The aim of this database is to provide researchers and scholars a unified database for potential data misreport by 171 countries regarding their COVID-19 daily reported cases. The analysis employs three different tests (chi-square, Kuiper, and Mean Absolute Deviation) to determine if the data given by each country in the world fit Benford’s Law.

## 1. Benford’s law

Benford’s Law (BL) is extensively used to test the legitimacy of data in numerous applications, such as in finance, economics, and politics. BL indicates that the first digit of a naturally transpire decimal number is more likely to be equal to 1, and the possibilities of the first digit to be equal to the subsequent numbers, decrease progressively.

The formula of BL is as follows, where *D* represents the digits between 1 and 9 and P is the probability:

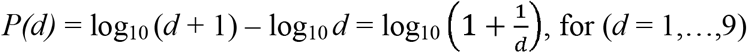

Using this formula, the expected frequencies for digits in first position are illustrated below:

| Digit | 1 | 2 | 3 | 4 | 5 | 6 | 7 | 8 | 9 |
| --- | --- | --- | --- | --- | --- | --- | --- | --- | --- |
| Expected Frequency | .30103 | .17609 | .12494 | .09691 | .07918 | .06695 | .05799 | .05115 | .04576 |

## 2. Tests

To measure the difference between the observed and expected first digit BL distribution for a country we used a *chi-square* test, which is based on the formula below:

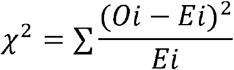

Where *Oi* is the observed and *Ei* is the expected absolute frequencies for digit *i*.

The *Kuiper* test (a modified Kolmogorov–Smirnov test) (Kuiper 1960) ignores sample size and it is based on the following formula:

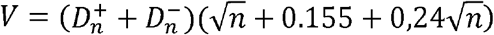

where 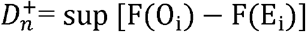, and 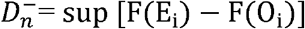 and F(.) stands for cumulated relative frequencies. The discrepancy statistics *D*^*+*^ and *D*^−^ characterize the absolute sizes of the differences between the two distributions being compared: the absolute and the observed distribution.

The *Mean Absolute Deviation (MAD)* test that also ignores sample size yet if the data sample size is small it may be inclined to false positives errors, when the results conclude nonconformity from unbiased data (Nigrini 2012). Henceforth, Nirgini’s MAD can only be applied to large sample size i.e. the total daily reported cases in the world. Johnson and Weggenmann (2013) have adjusted the MAD calculation method to better solve the false positive problem; hence, an adjusted MAD can be used for country’s data set. The MAD statistic is calculated as follows:

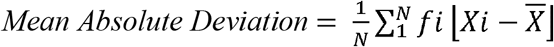

Where *Xi* is the difference between actual occurrence rate and Benford occurrence rate and 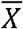 is the mean of the difference between actual occurrence rate and Benford occurrence rate. Whereas in Nigrini’s MAD, *Xi* is the actual occurrence rate and 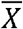 is the Benford occurrence rate. The absolute symbol means that the deviation is given a positive sign irrespective of whether it is positive or negative. Individual differences are then totaled and divided by 9 (the number of leading digits) to yield the mean absolute deviation (Johnson and Weggenmann 2013).

## 3. Data collection

We used publicly available data from the European Centre for Disease Prevention and Control website^1^ on reported daily cases of Covid-19 in the world starting from the first case of Covid-19 in each country to the 12^nd^ of November 2020.

The below Figure shows the Benfordness of the World’s reported daily cases where the blue bars represent the world’s observed daily reported cases and the intermittent red line represents BL.

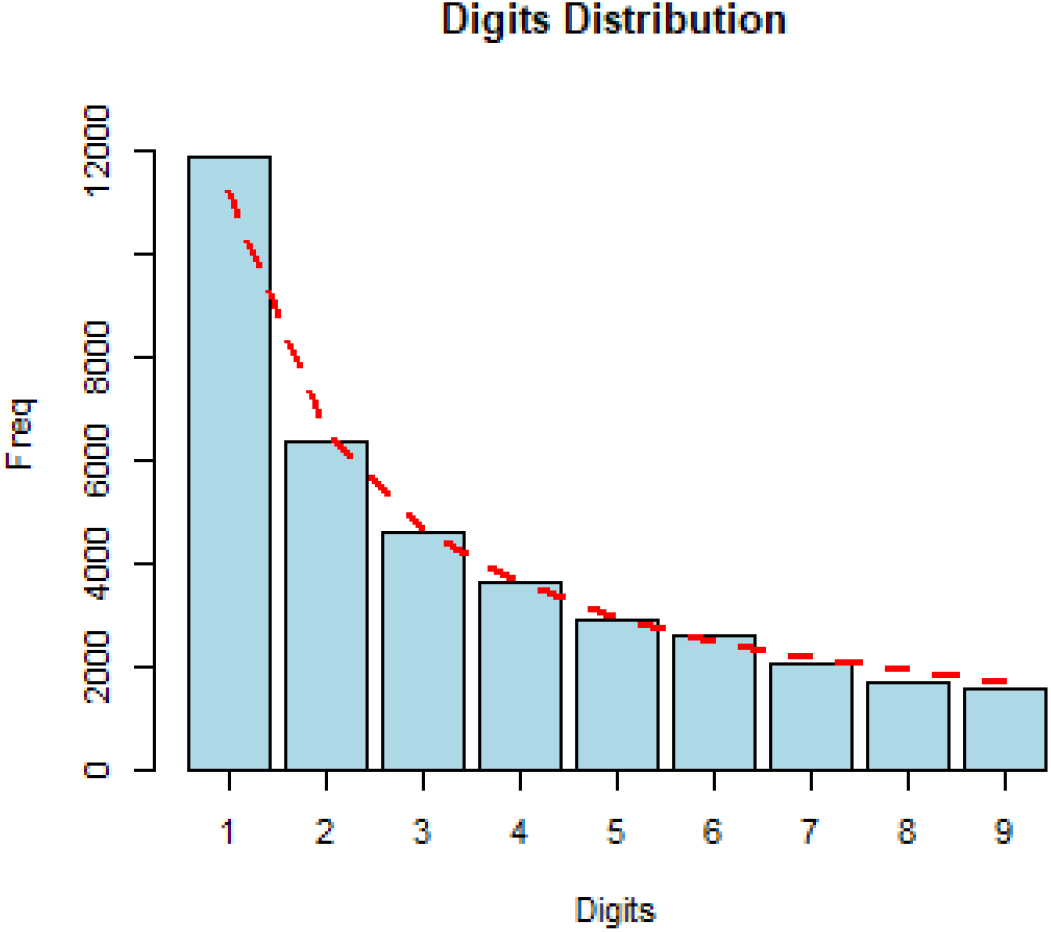

## 4. The database

The ranges of the tests are shown below. If the value of the chi-square and the Kuiper tests are more than 20.09 and 2 respectively, it is considered that a country’s distribution curve does not fit BL. Similarly, in the MAD test, if the value is more than 0.015, the curve does not conform to BL.

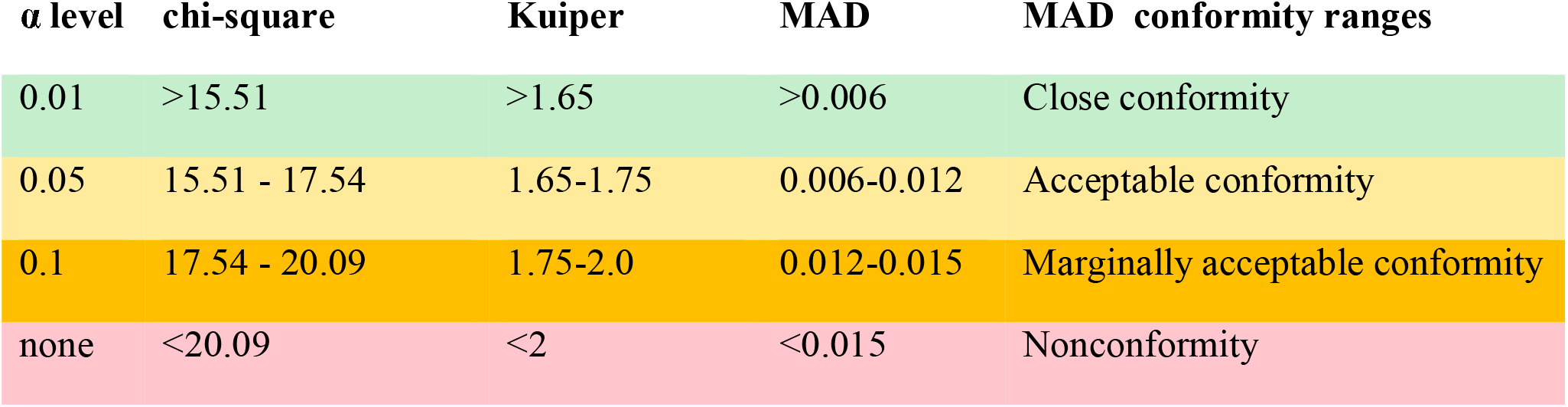
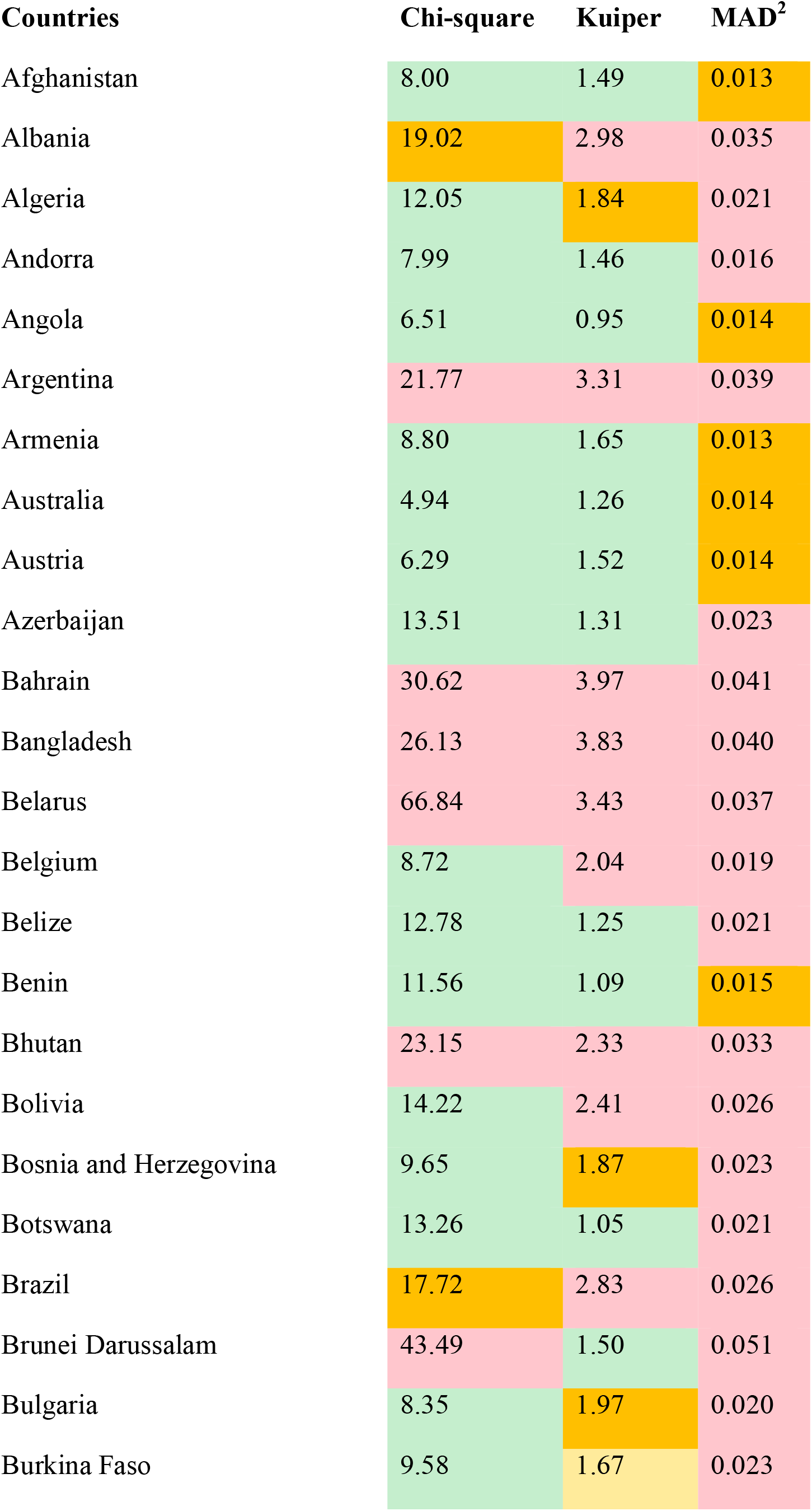
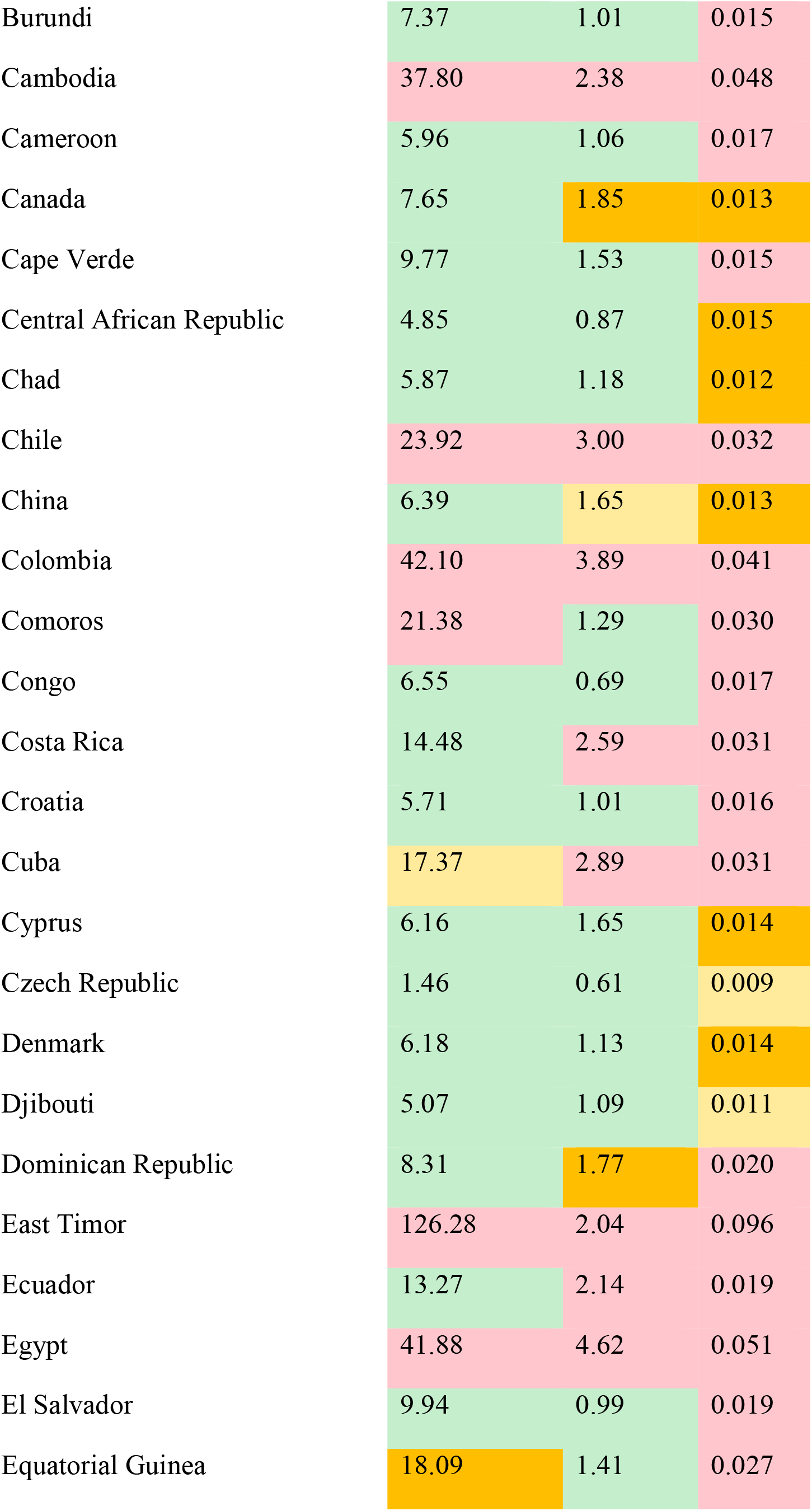
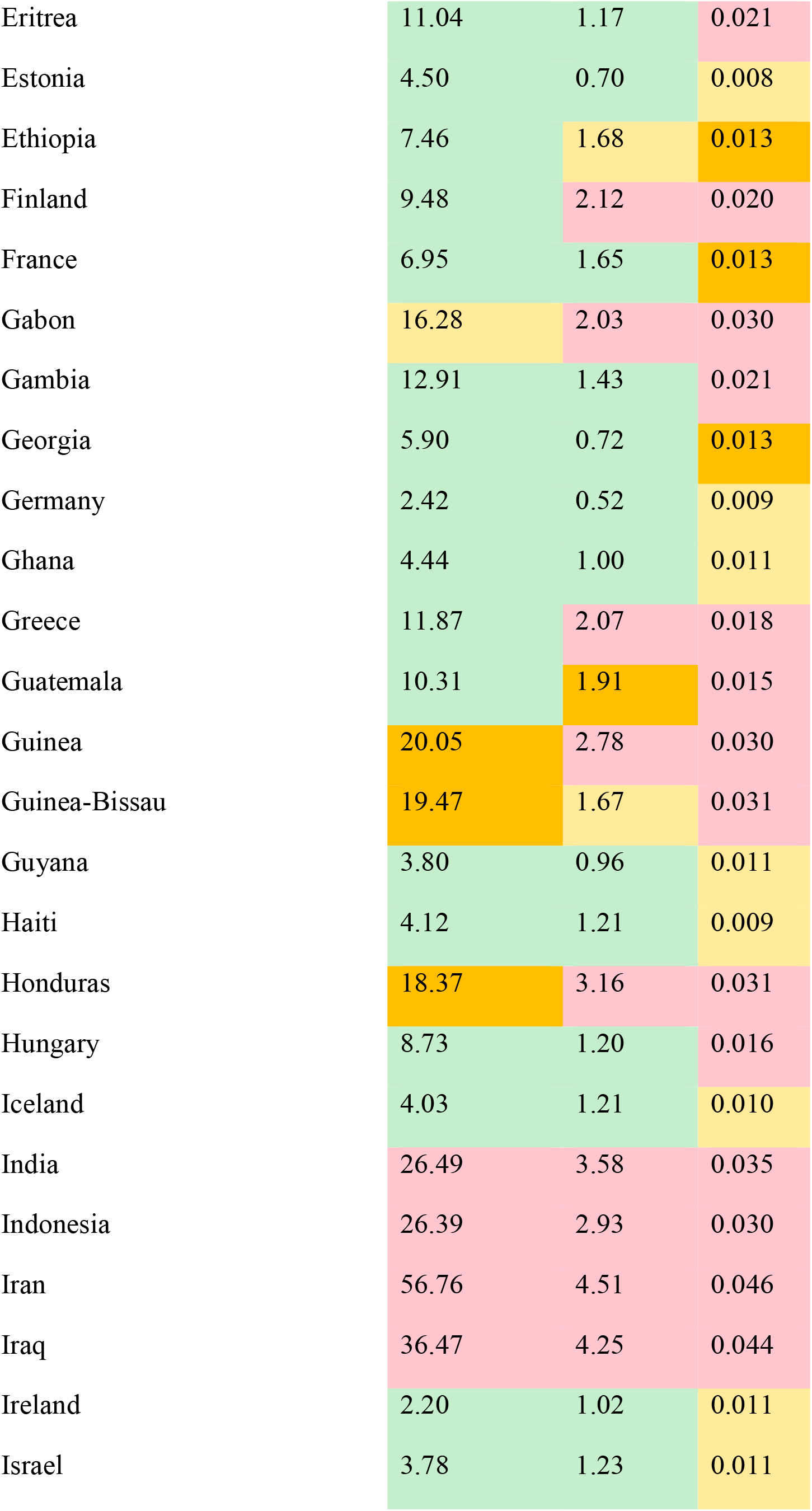
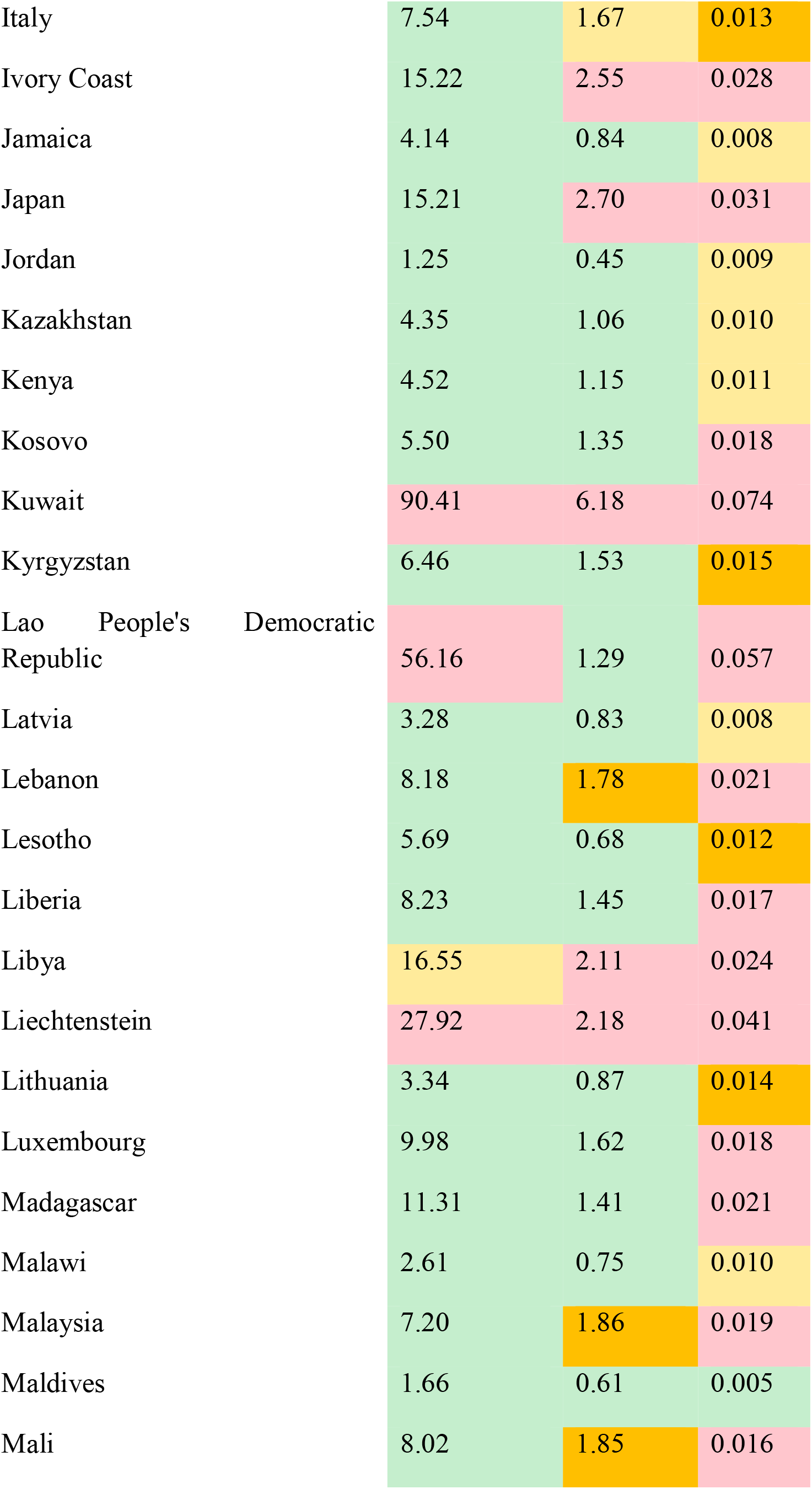
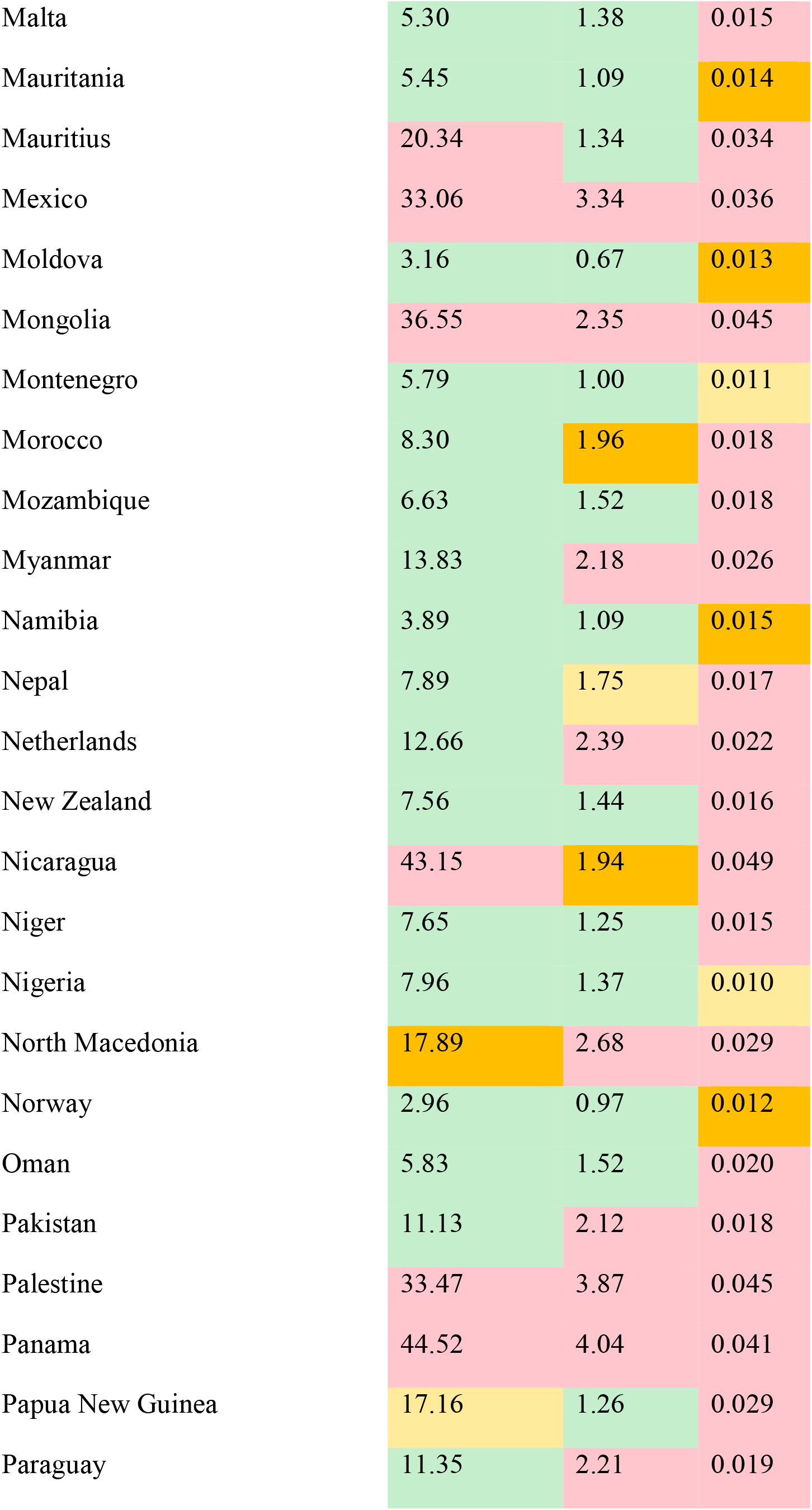
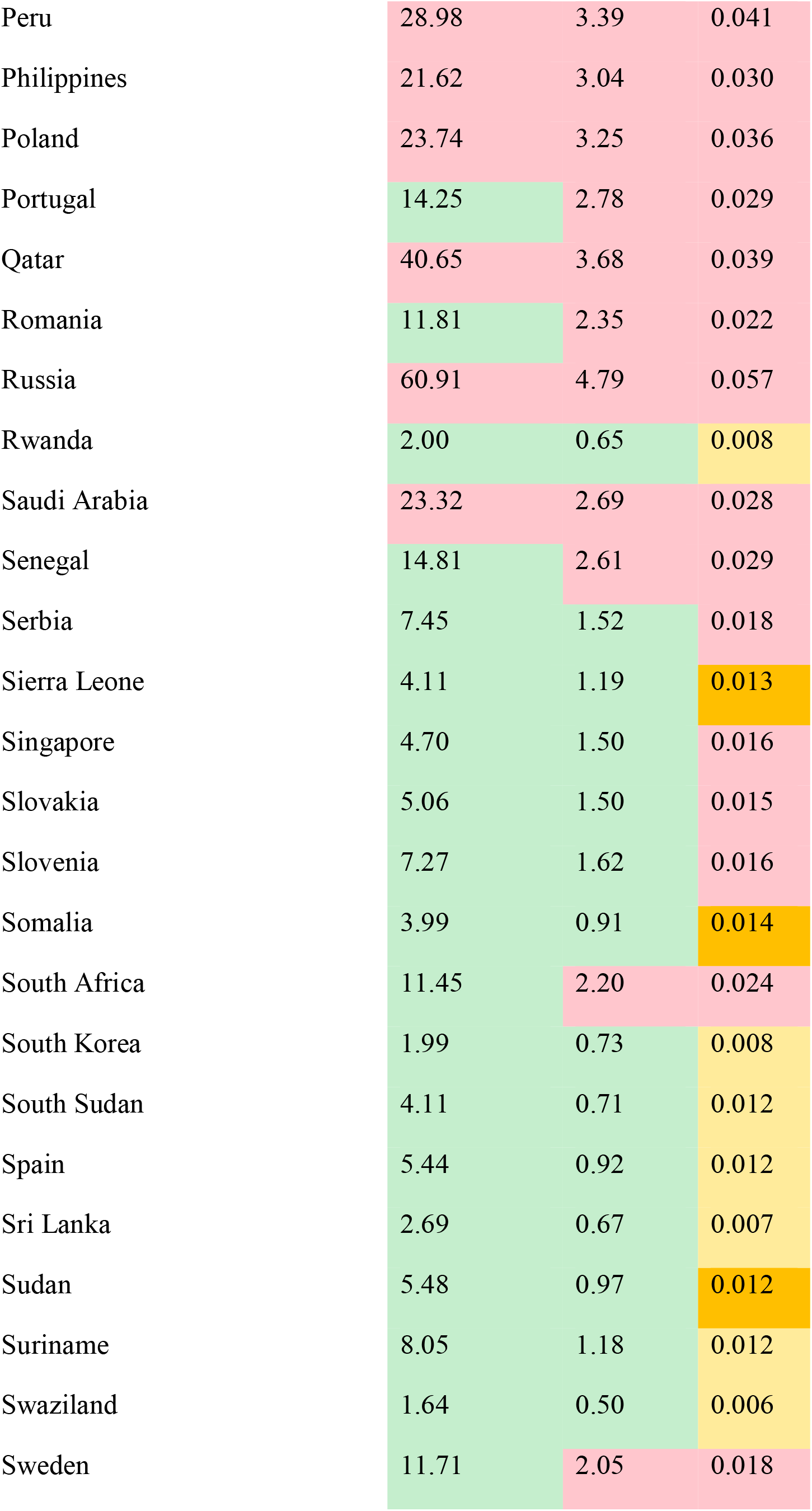
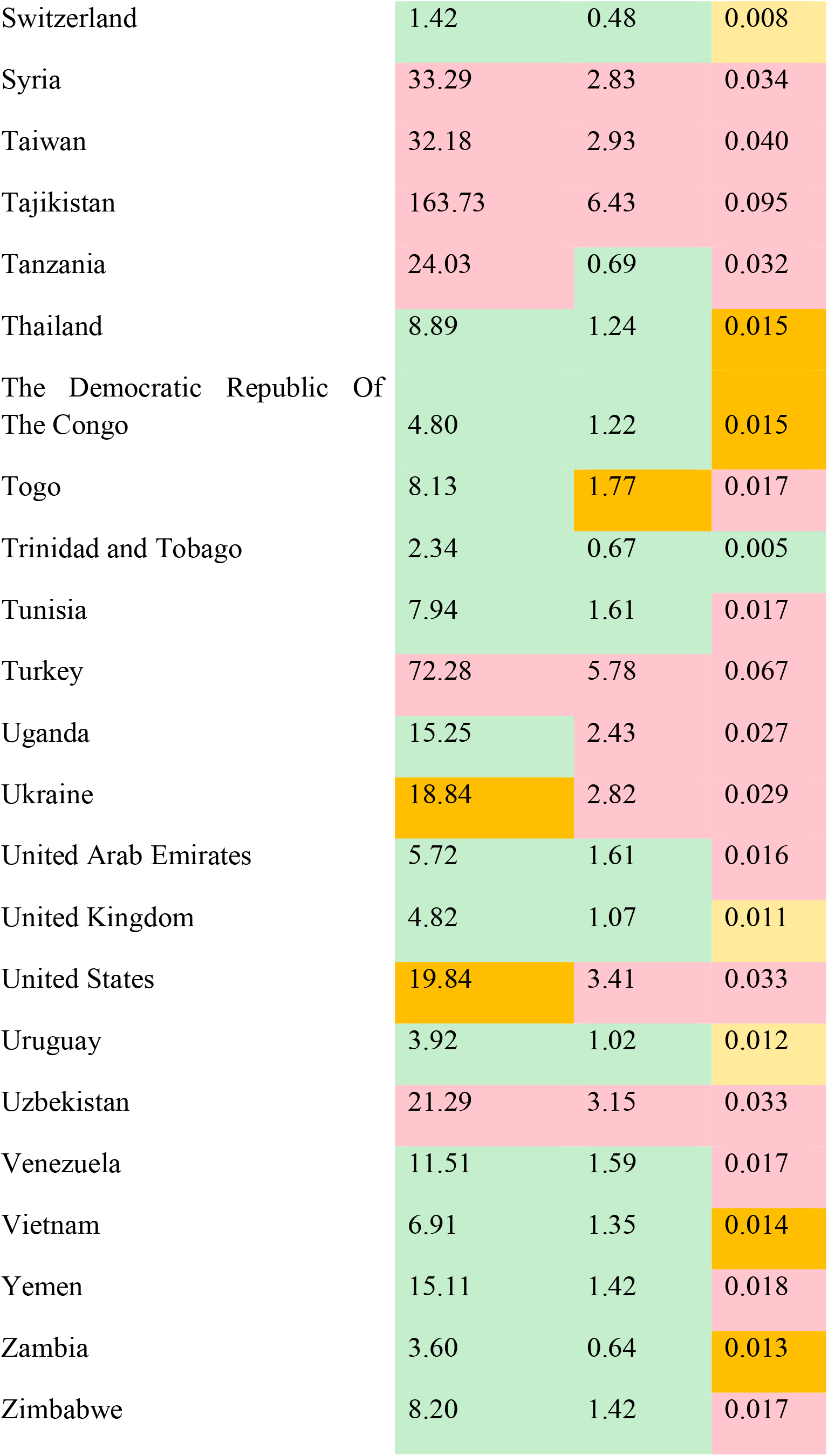

## Data Availability

The data is fully available in the paper.

## Footnotes

1 https://www.ecdc.europa.eu/en

